# TreadWill: Development and pragmatic randomized controlled trial of an unguided, computerized cognitive behavioral therapy intervention in a lower middle-income country

**DOI:** 10.1101/2021.11.24.21266799

**Authors:** Arka Ghosh, Rithwik J. Cherian, Surbhit Wagle, Parth Sharma, Karthikeyan R. Kannan, Alok Bajpai, Nitin Gupta

**Author notes:** Corresponding authors: Arka Ghosh and Nitin Gupta. Postal address: Dr. Nitin Gupta, Department of Biological Sciences and Bioengineering, Indian Institute of Technology Kanpur, Kanpur 208016, Uttar Pradesh, India.

## Abstract

Most individuals vulnerable to depression do not receive adequate or timely treatment globally. Unguided computerized cognitive behavioral therapy (cCBT) has the potential to bridge this treatment gap. However, the real-world effectiveness of unguided cCBT interventions, particularly in low- and middle-income countries (LMICs), remains inconclusive. In this study, we report the design and development of a new unguided cCBT intervention, TreadWill, and its pragmatic evaluation. TreadWill was designed to be fully automated, engaging, easy to use, and accessible in LMICs. To evaluate its effectiveness and engagement level, we performed a double-blind, fully-remote, randomized controlled trial with 598 participants in India. The use of TreadWill significantly reduced depression-related and anxiety-related symptoms. Compared with a plain-text version with the same therapeutic content, the full-featured version of TreadWill showed significantly higher engagement. Overall, our study provides a new resource and evidence for the use of unguided cCBT as a scalable intervention in LMICs.

## Introduction

More than 264 million individuals suffer from depressive disorders worldwide (James et al., 2018). Despite the availability of evidence-based pharmacological and psychological treatment approaches, 76% to 85% of individuals suffering from mental health disorders do not get any treatment in low- and middle-income countries (LMICs) (Wang et al., 2007). Barriers to accessing treatment for mental health disorders include the lack of access to treatment options, high cost, fear of social stigma, and an inclination to self-manage the problem (Andrade et al., 2014; Patel et al., 2018; Saxena et al., 2007). In India, there is a treatment gap of 85.2% for major depressive disorders (Gururaj et al., 2016).

One approach to bridge this treatment gap is to deliver computerized psychotherapy. The first therapeutic chatbot, ELIZA, was developed in 1966 (Weizenbaum, 1966); it was a rudimentary program based on text rephrasing rather than evidence-based methods. The first computer-assisted Cognitive Behavioral Therapy (CBT)-based program for depression was delivered in 1982 (Selmi et al., 1982). However, in the past two decades, the advent of stable internet connection and the pervasiveness of smartphones and computers have made it feasible to deploy technological interventions at scale.

Computerized CBT (cCBT) has gained traction as a viable treatment modality, with more than 200 trials conducted so far (Andersson & Carlbring, 2021). cCBT for depressive disorders, both guided and unguided, has been evaluated in several clinical trials worldwide. Recent studies and meta-analyses indicate that for depressive symptoms, guided cCBT interventions are more beneficial than unguided cCBT interventions (Cuijpers et al., 2019; Gilbody et al., 2017; Johansson & Andersson, 2012; Karyotaki et al., 2021; Wright et al., 2019). Carlbring et al. showed equivalent effects between guided cCBT interventions and face-to-face CBT (Carlbring et al., 2018). Including guided cCBT intervention with treatment-as-usual does not add any extra benefits (Gilbody et al., 2015). Moreover, while guided cCBT intervention can be a feasible option in developed countries (Andrews et al., 2018), it is not feasible in low- and middle-income countries (LMICs) due to the acute shortage of mental health professionals (Bruckner et al., 2011).

Unguided cCBT interventions have the potential to bridge the treatment gap in LMICs. The evidence for unguided cCBT interventions is mixed, with some meta-analyses showing that they are effective with a small or medium effect size (Karyotaki et al., 2017; Twomey et al., 2020; Wright et al., 2019) and some showing that they are not effective (Garrido et al., 2019; Harrer et al., 2019; Välimäki et al., 2017). Note that the unguided studies included in the meta-analyses often involved initial contact with humans for diagnostic interviews (Berger et al., 2011; de Graaf et al., 2009; Farrer et al., 2011; Meyer et al., 2015; Mira et al., 2017; Spek et al., 2007), weekly telephone contact support (Christensen et al., 2004; Phillips et al., 2014), or treatment-as-usual (Gilbody et al., 2015; Klein et al., 2016). Even minimal human contact can increase adherence to the interventions compared with a study without any such contact (Moritz et al., 2012; Morriss et al., 2021). Indeed, Fleming et al. (Fleming et al., 2018) found that adherence rates observed in trial settings fail to translate into the real world. Recent meta-analyses have reported a positive correlation between treatment adherence and treatment effects (Karyotaki et al., 2017; Wright et al., 2019). Also, a recent meta-analysis found that existing cCBT interventions, guided or unguided, had low acceptability among patients, less than even waitlist (Cuijpers et al., 2019).

Studies on cCBT interventions have been conducted predominantly in high-income countries (Lehtimaki et al., 2021), but see (Martínez et al., 2018) for depression and (Fu et al., 2020) for all mental health disorders in LMICs. A recent meta-analysis reported that 92% of the studies on diagnosed depression had been conducted in Western Europe, North America, or Australia (Köhnen et al., 2021). The interventions have been developed, evaluated, and made available for free only in these high-income regions. There is a need for unguided interventions that are more effective, have higher adherence, and are available free of cost for wide accessibility in LMICs.

In this study, we develop and evaluate such a cCBT intervention, TreadWill. We included several features in TreadWill that could increase adherence to and improve the effectiveness of a completely unguided intervention. We also developed an active control version of TreadWill that presented the same therapeutic content without said features. We designed a fully remote three-armed randomized controlled trial — experimental version of TreadWill, active control version of TreadWill, and waitlist control. We hypothesized that participants assigned to the experimental group would show significantly more improvement in depressive and anxiety symptom severity than the control groups. We also hypothesized that the participants in the experimental group would show significantly more engagement in terms of modules completed and absolute time used compared with the active control group participants.

## Methods

### Study design

We designed a fully remote randomized controlled trial to test the effectiveness of the experimental version of TreadWill compared with an active control version and a waitlist control version. We planned to recruit 600 participants in a 1:1:1 distribution across the three groups. We implemented simple randomization using an automated randomization function (developed in Python 3.4.3). This trial was registered in ClinicalTrials.gov before commencement (Identifier: NCT03445598)

### Participant recruitment and screening

We recruited participants using both offline and online publicity. We displayed flyers in the residential hostels, research buildings, and lecture halls of the Indian Institute of Technology (IIT), Kanpur. A press release helped with coverage in newspapers and social media.

The publicity material included a website link to join the study. The link opened a webpage that provided information about the study and accepted the email-id of the interested participants. Over the next three steps, the webpage collected the demographic data, the baseline Patient Health Questionnaire-9 (PHQ-9) score (Kroenke et al., 2001), and the informed consent from the potential participants. The entire participant recruitment process was automated to eliminate human contact. To be eligible to participate in the study, an individual must have been a 16–35-year-old Indian resident. They must have been fluent in English and must have had access to an internet-enabled computer or tablet device. They must have scored between 5-19 (both inclusive) in PHQ-9 with a score of 0 on the 9th question. We decided to include participants with mild symptoms of depression (a score of 5-9 in PHQ-9) and exclude those with severe symptoms (a score >19) because our program was targeted not at the clinical population but at a wider population with vulnerability for depression. We excluded individuals who were unemployed, had a diagnosis of bipolar disorder or psychosis, or reported that they just wanted to check out the site and did not plan to complete it (the last condition was added after trial commencement to exclude casual visitors to the website). Because of the pragmatic nature of our study, we included participants regardless of whether they were availing treatment for depression. Once a potential participant met the inclusion/exclusion criteria and provided informed consent (for individuals of age 16-17, informed consent was also required from a parent or guardian), the individual was scheduled to be recruited in the study. Eighteen hours later, the individual was randomized to one of the three groups and received a unique link via email. The delay of 18 hours was included to prevent individuals from signing up using disposable temporary email ids. They were counted as a participant in the study only after clicking on the unique link and were led to a signup page (for participants assigned to the experimental or active control group). The participants assigned to the waitlist control group were led to a page to collect their baseline Generalized Anxiety Disorder-7 (GAD-7) score (Spitzer et al., 2006); GAD-7 scores of experimental and active control groups were taken as part of the intervention. Participants did not receive any monetary compensation.

### Ethics

The Institutional Ethics Committee, Indian Institute of Technology Kanpur, provided the ethical clearance to conduct this study (IITK/IEC/2017-18 II/1).

### Safety check

At any stage in the intervention, if we detected severe depressive symptoms or suicidal ideation, we blocked access to TreadWill. Severe depressive symptoms were determined as a total score >19 in PHQ-9. Suicidal ideation was detected by a score >0 in the 9th question of PHQ-9 and a total score >4 in the Suicidal Intent Questionnaire (SIQ) (Gupta et al., 1983). In such cases, an email and SMS alerts were sent to the participants (and their *buddy*, if they had one in the program) requesting them to seek professional help. For participants aged 16-17 years, an email notification was sent to the parent or guardian as well.

### Automated notifications

Participant contact was minimal and automated. Participants who initiated the recruitment process but did not complete it were sent automated email reminders encouraging them to complete the process. Participants also received periodic automated email and SMS reminders nudging them to use TreadWill (see Supplementary Table S1 for more details). The program asked the participants about their preferred time to log in; using this information, email and SMS alerts were sent 10 minutes in advance to remind the participants. The research team did not initiate any direct contact with the participants. Technical support via email was provided in case the participants sent an email requesting it.

### Development of the intervention

We used the Django framework for developing the TreadWill website. We used Google Slides to embed the slides and YouTube to embed the videos in the website. We used images with a Creative Commons license for use in Slides and Videos. We used images from pics.stir.ac.uk for the “Identify the friendly face” game. The content and the website went through multiple rounds of checking by the development team and other volunteers to fix errors before launching the trial.

### Assessments

We used Patient Health Questionnaire-9 (PHQ-9) (Kroenke et al., 2001) and Generalized Anxiety Disorder-7 (GAD-7) (Spitzer et al., 2006) questionnaires to measure depressive and anxiety symptom severity, respectively.

For experimental and active control group participants, PHQ-9 was administered at baseline, at the beginning of each module (*except* the first module), after completing all the modules, and at a 90-day follow-up. GAD-7 was administered at the beginning of each module (*including* the first module), after completing all the modules, and at a 90-day follow-up. For the waitlist control group, PHQ-9 and GAD-7 were administered at baseline and after a 42-day interval (this interval was chosen to be at par with the expected intervention duration of approximately 6 weeks for completing the 6 modules in the experimental group). After the 42-day interval, the waitlisted participants were also given access to the intervention.

### Blinding

All participants followed the same recruitment procedure. Consequently, the participants were unaware of which version of TreadWill they were assigned to (they did not even know that two different versions existed). Therefore, we expect the placebo effects in the two groups to be similar. Also, the PHQ-9 and GAD-7 data were self-reported on the website, so there was no scope for evaluator bias.

### Statistical analyses

Due to the high dropout rate, we did not assume the PHQ-9 and GAD-7 scores to be normally distributed and therefore used non-parametric statistical tests for analyzing the effectiveness of the intervention. We used the Kruskal-Wallis test for comparing the reduction in depression or anxiety symptom severity from baseline to the primary endpoint between the three groups. For post-hoc analysis between the groups, we used Mann-Whitney U test. The tests were implemented in MATLAB and Python.

Since this was the first trial of TreadWill, we did not have a prior estimate of the dropout rate and could not do power calculations. We chose the sample size of 600 participants based on previous studies of similar nature (Arean et al., 2016; Gilbody et al., 2015).

## Results

### Approach taken for developing the intervention

We aimed to develop and evaluate a fully automated cCBT intervention, TreadWill, that would be engaging and effective without any expert guidance or contact. We surveyed the existing cCBT interventions before starting the development process and considered factors that may be responsible for the high dropout rates. The common shortcomings that we identified included lack of interactive content, lack of tailoring of the content to different users, lack of peer support for users, and lack of engaging games. Different interventions addressed some of these shortcomings by including the corresponding features, but none of them included all the features. We developed TreadWill to address these shortcomings and hypothesized that it would lead to a high adherence rate and a significant reduction in depressive and anxiety symptom severity. As we do not plan to charge the users, we also expect TreadWill to be more accessible, especially in LMICs, compared to the paid interventions.

### Design of TreadWill

We designed the therapeutic content of TreadWill based on Cognitive Behavioral Therapy (CBT), using Beck 2011 as the primary reference (Beck, 2011). TreadWill delivers the core concepts of CBT in a structured format with six modules (see Supplementary Table S2 for details), in easily understandable language. Each module consisted of four sections – *Introduction, Learn, Discuss*, and *Practice*. In the *Introduction* section, an automated virtual therapist explained the importance of the module through interactive text-based dialog. The *Learn* section included psychoeducation in the form of slides and videos. Slides consisted of multiple infographics presented sequentially (Supplementary Fig. S1). Videos consisted of animated content with a voiceover explaining the concepts visible on the screen. In the *Discuss* section, the participant learnt to apply the psychoeducation to real-life situations through *conversations*. These *conversations* were text-based dialogues with an automated virtual patient (Supplementary Fig. S2) presented in an interactive format designed to simulate human chat. Although the conversations were pre-programmed, in many places the participants could choose from more than one response, thus providing some control to the user in steering the dialog. The *Practice* section included interactive quizzes on the material covered in each module.

To ensure sequential progression through the intervention, only the first module was initially accessible to the user and the later modules were locked. After completion of all sections in a module and a 4-day gap since its unlocking (to prevent rushing through the modules), the next module was unlocked. Steps within a module were also unlocked gradually upon completion of preceding steps. Each participant had a maximum of 90 days to complete the six modules starting from their first login. After 90 days, they could continue to use the modules unlocked until then but could not unlock new modules. We did this to restrict participants’ exposure to new therapeutic content after 90 days and provide a clear deadline, as recommended by several studies (Andersson et al., 2009; Meyer et al., 2009; Nordin et al., 2010).

### Interactive games and CBT forms in TreadWill

TreadWill included two interactive games. The “Identifying thinking errors” game was aimed at training the participants in spotting thinking errors in their negative automatic thoughts. The gameplay involved presentation of a situation, a related negative automatic thought, and a list of 10 thinking errors, from which the participant had to select one or more thinking errors present in the thought. Selecting a correct option allowed the participant to move to the next level. On selecting an incorrect option, feedback was provided along with an opportunity to try again. The “Identify the friendly face” game is based on the training paradigm developed by Dandeneau and Baldwin to train participants to overcome the negative attention bias and improve their self-esteem (Dandeneau & Baldwin, 2009), thereby reducing the risk of depression (Orth et al., 2016; Sowislo & Orth, 2013). The game presented 4 images in a 2×2 grid with 3 faces showing a negative emotion and 1 face showing a positive emotion. The participant was allowed 5 seconds to find the positive image and thus increase their score. If the participant responded or if 5 seconds elapsed, a new set of images was shown. The gameplay incentivized quick attention to positive emotions. The difficulty of the game continuously adapted to the participant’s competence: incorrect responses increased the frequency of faces with obvious emotions and correct responses increased the frequency of faces with subtle emotions.

TreadWill provided an interactive interface to fill the forms commonly used in CBT: *Thought record worksheet, Core belief worksheet, Behavioral experiment worksheet, Problem solving worksheet, Prepare for setback worksheet, and Schedule activity worksheet* (Supplementary Table S3). The forms allowed participants to apply the techniques of CBT to their situations and save the information for future reference.

### Peer and family support in TreadWill

Individuals looking for support online might have low social support in real life (Houston et al., 2002). In such cases, online peer-based support has been shown to be effective in reducing depressive symptoms (Griffiths et al., 2012; Tomasino et al., 2017). Keeping this in mind, we designed the *SupportGroup* and *PeerGroup* features in TreadWill to provide a social space where participants could connect with other TreadWill users and potentially help each other in solving their problems. Posts in the *SupportGroup* were visible to all TreadWill participants. The participants could upvote or downvote posts, add comments, and send “thank you” messages to each other. *PeerGroups* were smaller groups of 10 members each, designed such that the posts in a *PeerGroup* were visible only to the members of that *PeerGroup*.

We provided the participants an option to invite a family member or a friend as their *buddy* who would receive weekly updates about the participant’s activities in TreadWill. We hypothesized that the involvement of the *buddy* would motivate the participant to complete the program. We sent an email to this *buddy* if the participant failed to use TreadWill regularly and requested them to nudge the participant.

### Content tailoring in TreadWill

Content tailoring has the potential to increase adherence to cCBT interventions as participants are more likely to stick with a program if they find the content relatable (Carlbring et al., 2011; Johansson et al., 2012; Meyer et al., 2009). In TreadWill, we implemented tailoring by selecting the examples in the *conversations* based on the participant’s occupation (high-school student, college student, or working professional). In addition, we also tailored the *conversations* based on the participant’s thoughts, beliefs, and situations in the following way. First, we asked the participant to select relatable intermediate and core beliefs from the Dysfunctional Attitude Scale (Weissman & Beck, 1978), negative automatic thoughts from the Automatic Thoughts Questionnaire (Hollon & Kendall, 1980), and stressful situations from a curated list. Then, we made the simulated virtual patients in the subsequent *conversations* identify with similar beliefs, thoughts, and situations, and the participant’s goal was to help the simulated patient using the techniques of CBT learnt in that module.

The automated email and SMS notifications received by the users were also tailored according to their preferences (see Methods).

### Active control version

The active control version presented the same CBT content as the experimental version, in the same six modules, but using plain text instead of slides, videos, and conversations. Each module had *Introduction, Learn*, and *Discuss* sections, but the *Practice* section was excluded. The content was not tailored according to the participant. The active control version included the CBT forms, but excluded games, *SupportGroup, PeerGroup*, and the option to involve a buddy. The participants received only essential email notifications (see Supplementary Table S1 for details of notifications).

### Participants

Recruitment commenced on February 14, 2018, with a planned enrolment of 600 participants. The primary completion date was March 2, 2019, after full enrollment, and the secondary completion date was May 31, 2019. Out of the 5188 individuals who started the registration process for the study, 598 participants completed all the steps and met the study inclusion criteria (2 other participants who did not meet the inclusion criteria were initially included due to a software bug but were excluded when we cross-checked the data during data analysis). The 598 participants were randomly assigned to the three study arms with equal probability (see Methods), resulting in 204 as experimental (EXP), 189 as active control (ACT), and 205 as waitlist control (WL) participants (Fig. 1). The participants in the three groups were found to be balanced in terms of age, gender, the severity of depressive symptoms, occupation, use of other interventions, motivation for joining, and occurrence of recent traumatic events (Table 1). However, a gender bias (79% male) was observed because participants in our study were recruited mainly from Indian engineering colleges where the students are predominantly male (*AISHE Final Report, 2018-19*). Also, in India, there is a 56% gender gap in mobile internet usage (*The Mobile Gender Gap Report 2019*).

**Figure 1:**
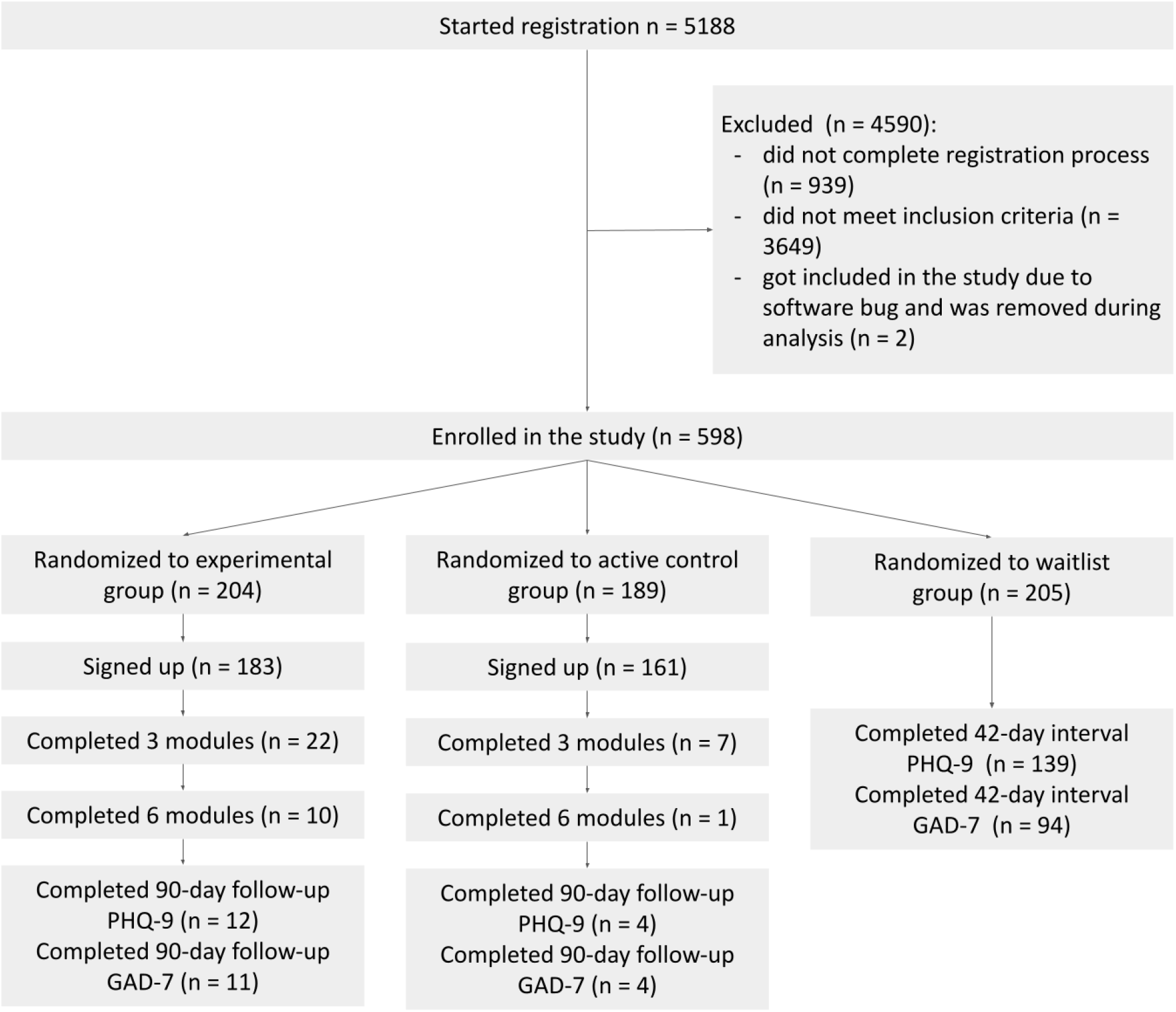
The flow of participants in the trial. In the experimental and the active control groups, the follow-up scores of only those participants were analyzed who had completed at least 3 modules. In the waitlist group, the 42-day interval scores of only those participants were analyzed who had also submitted the baseline scores.

**Table 1:** Baseline and demographic characteristics of the participants recruited in the study.

| **Group (n)** | **Experimental**<br>**(n=204)** | **Active**<br>**Control**<br>**(n=189)** | **Waitlist**<br>**Control**<br>**(n=205)** | **All (n=598)** |
| --- | --- | --- | --- | --- |
| **Age**, Mean (s.e.) | 23.76 (0.30) | 23.42 (0.28) | 23.48 (0.29) | 23.56 (0.17) |
| **Gender**, (male, female) | (160, 44) | (151, 38) | (167, 38) | (478, 120) |
| **Traumatic event or death of a loved one**, (Yes, No) | (22, 182) | (18, 171) | (20, 185) | (60, 538) |
| **Joining for help**, (Yes, No) | (180, 24) | (157, 32) | (180, 25) | (517, 81) |
| **Secondary help**, (None, Counselling, Medication, Both) | (185, 5, 12, 2) | (178, 4, 4, 3) | (193, 4, 6, 2) | (556, 13, 22, 7) |
| **Occupation**, (High School Student, Between school and college, College student, Coaching after college, Working professionals, Self employed, Freelancers, Volunteers) | (1, 8, 113, 33, 39, 7, 2, 1) | (2, 4, 103, 33, 40, 4, 3, 0) | (4, 6, 120, 30, 37, 4, 2, 2) | (7, 18, 336, 96, 116, 15, 7, 3) |
| **PHQ-9**, Mean (s.e.) | 10.76 (0.26) | 10.81 (0.25) | 10.42 (0.27) | 10.66 (0.15) |

### Effectiveness of TreadWill

In the primary analysis, we included the participants who completed at least three modules in the experimental group (EXP) or the active control group (ACT). For this analysis, we used the last PHQ-9 and GAD-7 scores submitted by these participants excluding the follow-up questionnaire. Henceforth, we refer to the time of these last scores as the primary endpoint. From the waitlist control group (WL), all users who submitted the questionnaires after the waiting period were included in the analysis.

We compared the reductions in the PHQ-9 and GAD-7 scores from the baseline to the primary endpoint between the three groups (Fig. 2A, B; Table 2). The three groups showed significant differences in the reductions in the PHQ-9 score (H(2) = 8.93, p = 0.012, Kruskal-Wallis test); a post-hoc test with Bonferroni correction revealed that the experimental group showed a larger reduction than the waitlist control group (2.73 versus 1.12, Mann-Whitney U = 1027, n_EXP_ = 22, n_WL_ = 139, p = 0.039). Similarly, the three groups showed significant differences in the reductions in the GAD-7 score (H(2) = 8.02, p = 0.018, Kruskal-Wallis test); a post-hoc test with Bonferroni correction showed a larger reduction in the experimental group than the waitlist control group (3.27 versus 0.89, Mann-Whitney U = 637.50, n_EXP_ = 22, n_WL_ = 94, p = 0.015). The differences in PHQ-9 or GAD-7 reductions between the experimental and the active control groups were not significant (PHQ-9: U = 96, p = 0.34; GAD-7: U = 52.5, p = 0.22; n_EXP_ = 22, n_ACT_= 7).

**Figure 2:**
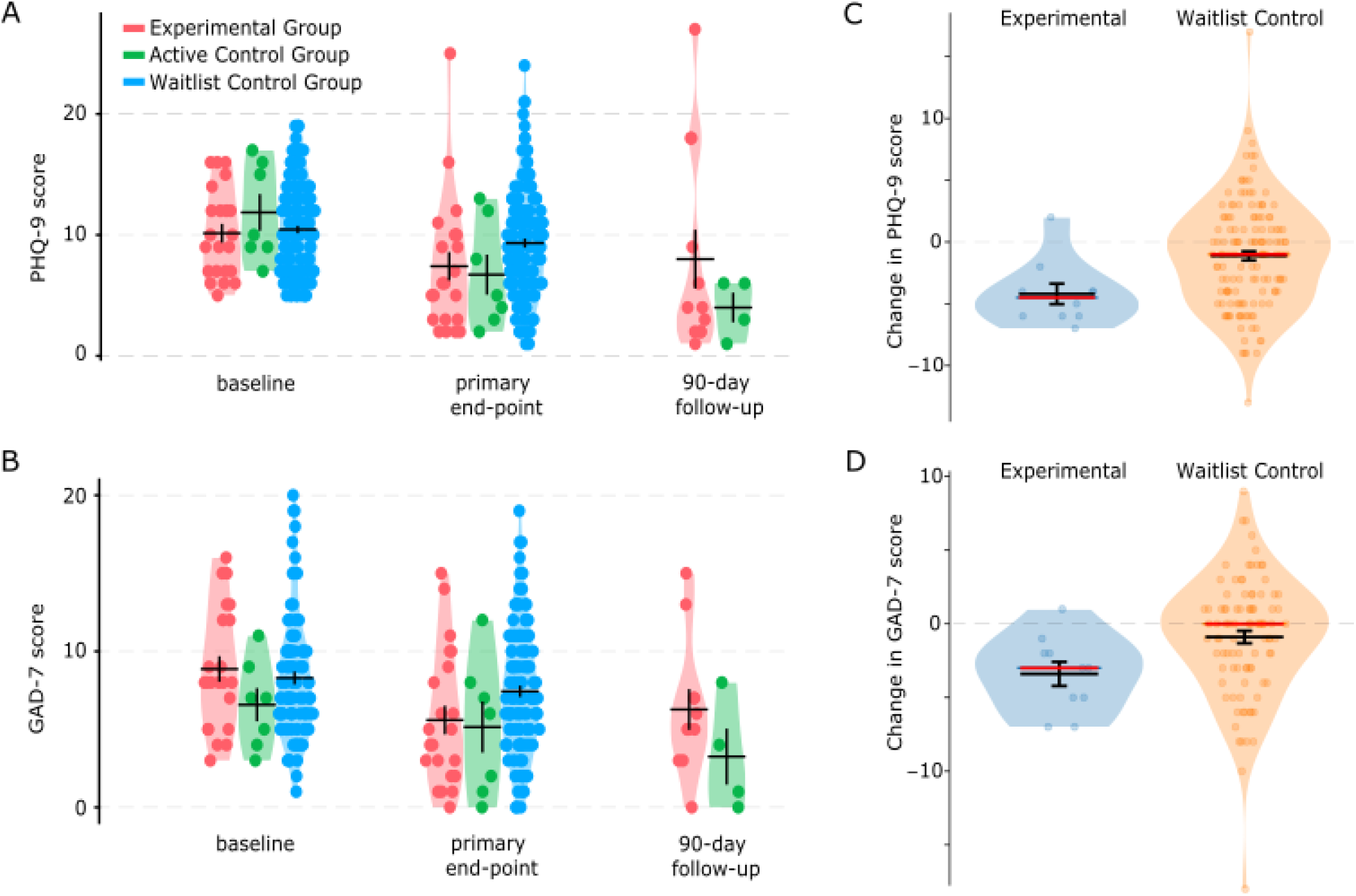
Changes in PHQ-9 and GAD-7 scores after using TreadWill. **A**,**B**, Violin plots show PHQ-9 (**A**) or GAD-7 (**B**) scores at baseline, primary endpoint, and follow-up for the experimental group, the active control group, and the waitlist group participants. Primary endpoint is defined as the latest PHQ-9 or GAD-7 score submitted after completing three modules. **C**,**D**, Violin plots show the change from baseline to program completion in PHQ-9 (**C**) or GAD-7 (**D**) score for the experimental group participants who completed all 6 modules (blue violin). For waitlist group participants (orange violin), the plots show the change from the score at the baseline to the score after the 42-day waiting interval (considered as the primary endpoint for the waitlist group). Red horizontal lines: median, black: mean. Error bars represent s.e.

**Table 2:** Average changes (± s.e.) in PHQ-9 and GAD-7 scores for the three groups from the baseline to the primary endpoint or to the completion of all modules. As only 1 participant in the active control group completed all modules, the corresponding values were not analyzed.

|  | **Experimental** | **Active control** | **Waitlist control** |
| --- | --- | --- | --- |
| PHQ-9 (primary endpoint - baseline) | -2.73 ± 1.27(n=22) | -5.14 ± 2.28 (n=7) | -1.12 ± 0.37 (n=139) |
| PHQ-9 (completion of all modules - baseline) | -4.20 ± 0.83 (n=10) | - |  |
| GAD-7 (primary endpoint - baseline) | -3.27 ± 0.97 (n=22) | -1.43 ± 0.92 (n=7) | -0.89 ± 0.42 (n=94) |
| GAD-7 (completion of all modules - baseline) | -3.40 ± 0.82 (n=10) | - |  |

We also checked the reduction in the PHQ-9 and the GAD-7 scores for the smaller set of the experimental group participants who completed all six modules (Fig. 2C, D; Table 2); this analysis could not be performed for the active control group because only 1 participant from that group completed all six modules. This analysis also showed that the experimental group had a significantly larger reduction in PHQ-9 scores compared to the waitlist control group (4.20 versus 1.12, Mann-Whitney U = 368.5, n_EXP_ = 10, n_WL_ = 139, p = 0.013) and GAD-7 scores (3.40 versus 0.89, Mann-Whitney U = 260.5, n_EXP_ = 10, n_WL_ = 94, p = 0.021). The 11 participants who completed all modules in the experimental and the active control groups did not differ demographically or in the baseline PHQ-9 score from the rest of the participants (Supplementary Table S5).

The reductions observed in the PHQ-9 and GAD-7 scores in the experimental and the active control groups at the primary endpoint were maintained at the 90-day follow-up (Fig. 2A, B). Thus, both the full-featured version of TreadWill (experimental) and the plain-text version of TreadWill (active control) were effective in reducing depression-related and anxiety-related symptoms of the participants who completed all or at least 3 modules.

We checked if the novel features of the experimental version of TreadWill were able to increase engagement compared to the active control version. Every module was completed by more participants in the experimental version than the active control version (Fig. 3A). The odds of completing at least 3 modules were three times larger for a participant in the experimental group compared to a participant in the active control group (odds ratio = 3.004; p = 0.011; 95% confidence interval = [1.247, 7.237]). The experimental group participants used TreadWill for an average of 79.8 minutes and the active control group participants for 26.1 minutes; the difference was statistically significant (Mann-Whitney U = 10290, n_EXP_=181, n_ACT_=159, p = 5.8 × 10^−6^, Fig. 3B). Thus, the full-featured version of TreadWill had higher engagement and less attrition than the plain-text version.

**Figure 3:**
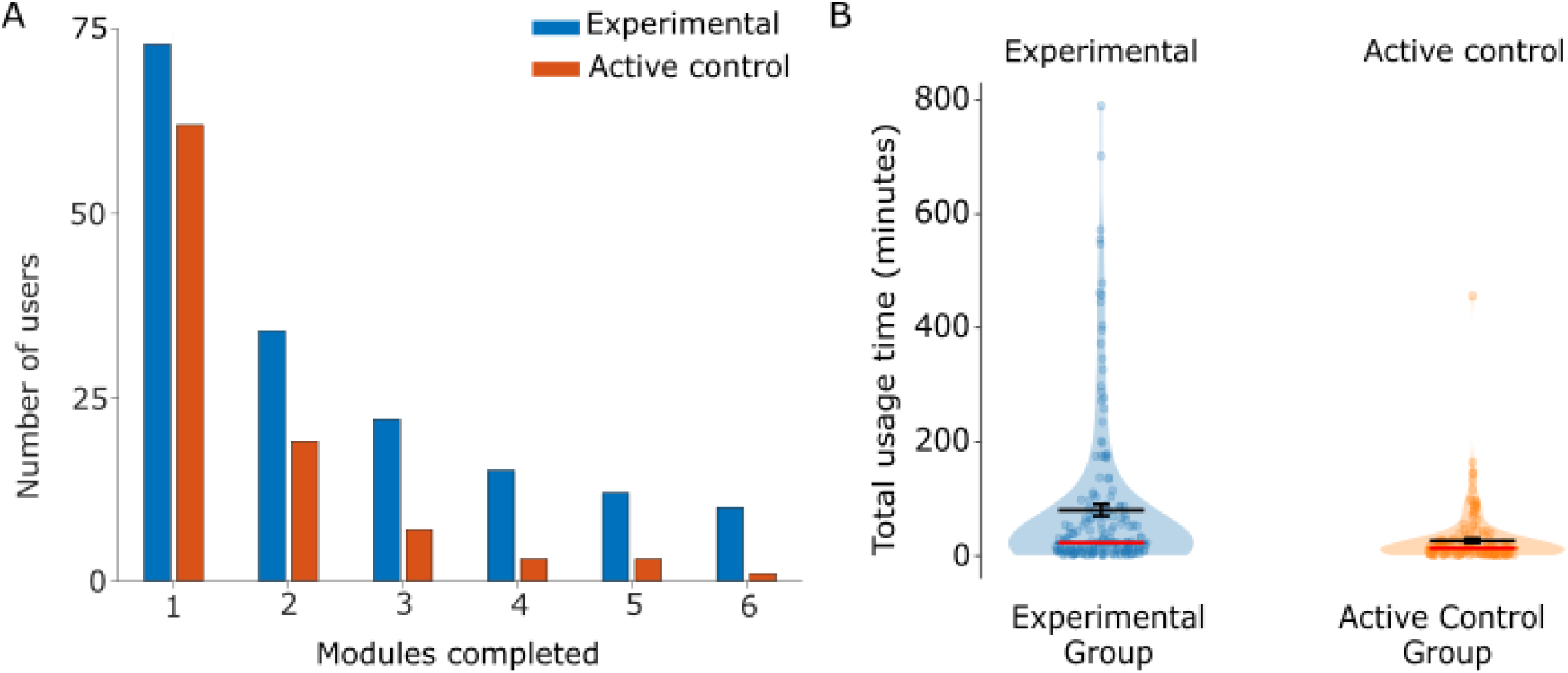
Adherence with TreadWill. **A**, The bar graph shows the number of participants in the experimental and the active control groups who completed the indicated number of modules. **B**, Violin plots show total usage times of the experimental and the active control group participants. Red horizontal lines: median, black: mean. Error bars represent s.e.

Further, we checked if the level of engagement with TreadWill was related to the reductions in depressive and anxiety symptoms. We found that the reduction in the PHQ-9 score was positively correlated with the number of modules completed (experimental group: Spearman ρ = 0.38, p = 0.003, n = 61, Sup. Fig. S3A; active control group: ρ = 0.51, p = 6.8 × 10^−4^, n = 41, Sup. Fig. S3B) and with the total usage time (experimental group: ρ = 0.39, p = 0.002, n = 61, Sup. Fig. S3C; active control group: ρ = 0.47, p = 0.002, n = 41, Sup. Fig. S3D). The reduction in the GAD-7 score was also moderately correlated with the number of modules completed (experimental group: ρ = 0.27, p = 0.039, n = 57, Sup. Fig. S4A; active control group: ρ = 0.43, p = 0.009, n = 37, Sup. Fig. S4B) and with the total usage time (experimental group: ρ = 0.25, p = 0.065, n = 57, Sup. Fig. S4C; active control group: ρ = 0.35, p = 0.035, n = 37, Sup. Fig. S4D).

### Feedback on the features of TreadWill

We programmed TreadWill to present surveys containing 15 questions using a 5-point Likert scale to quantify the participants’ feedback on various aspects of TreadWill. For example, one of the questions stated “I found the email reminders helpful”, to which the participant responded by selecting one of the following options: “strongly agree”, “somewhat agree”, “neither agree nor disagree”, “somewhat disagree”, and “strongly disagree”, which were mapped to a score of 2, 1, 0, -1, and -2, respectively (see Supplementary Table S4 for all questions). The surveys were conducted at two times points: after completing three modules and at the end of the intervention.

In the experimental group, the first survey was submitted by 22 participants and the second survey by 18 participants who completed at least 3 modules. The participants reported positive feedback on most aspects of TreadWill (Fig. 4A): mean feedback scores over all questions were significantly greater than zero for both the first survey (1.16 ± 0.12, n = 15 questions, t = 9.20, p = 2.60 × 10^−7^, t-test) and the second survey (1.29 ± 0.10, n = 15 questions, t = 12.56, p = 5.21 × 10^−9^, t-test). The scores remained largely consistent between the two surveys (Pearson r = 0.87, p = 2.84 × 10^−5^, n = 15). Strongest positive feedback was received for questions related to the ease of English used (1.86 ± 0.10 in the first survey & 1.83 ± 0.12 in the second survey), the relatability of the examples (1.23 ± 0.25 & 1.72 ± 0.13), the ease of using the CBT forms (1.45 ± 0.18 & 1.22 ± 0.17), the engaging nature of the conversations (1.36 ± 0.21 & 1.50 ± 0.20), the helpfulness of the *Learning* slides (1.73 ± 0.10 & 1.67 ± 0.14), and the helpfulness of the *Learning* videos (1.55 ± 0.13 & 1.67 ± 0.14). The features with the lowest ratings included the *PeerGroup*, which received weak positive feedback (0.73 ± 0.23 & 0.61 ± 0.28) and the *buddy* feature, which received neutral feedback in both surveys (−0.09 ± 0.22 & 0.39 ± 0.20).

**Figure 4:**
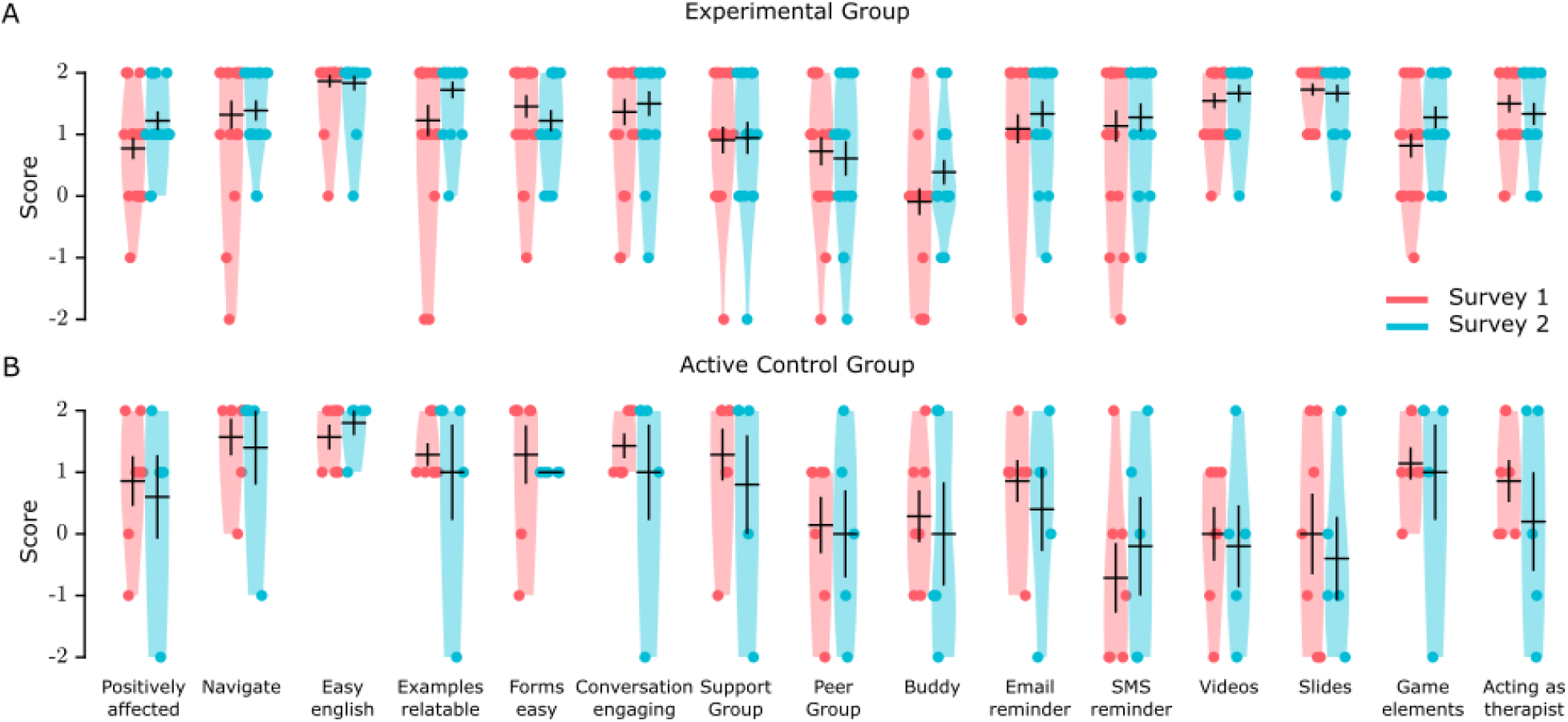
Feedback on the features of TreadWill. Violin plots show survey responses by the experimental group (**A**) and the active control group participants (**B**). The full texts of the survey questions are listed in Supplementary Table S4. Y-axis labels: 2 = “strongly agree”, 1 = “somewhat agree”, 0 = “neither agree nor disagree”, -1 = “somewhat disagree”, -2 = “strongly disagree”. Black lines indicate mean ± s.e.

In the active control group, only 7 and 5 participants who completed at least 3 modules submitted the first and the second survey, respectively. The survey questions were slightly different in the active control group (Supplementary Table S4): the first 6 questions judged their opinion on aspects they experienced directly, and the next 9 questions were asked in a prospective manner; for example, “I would prefer to have email reminders”. In the first 6 questions, the participants reported overall positive feedback (Fig. 4B). In the 9 prospective questions, they showed interest in having only some of the proposed features, including conversations and game elements. Curiously, many features that the active control group participants thought they would not prefer – including SMS reminders, videos, and slides – were actually found to be useful by the experimental group participants who experienced the features (Fig. 4A, B).

No participant reported any adverse events through the contact form on the website.

### Exploratory analysis: Feedback on the content

To check if the content provided was engaging, we provided the experimental group participants the option to provide feedback on the slides, videos, and *conversations* using *like* and *dislike* buttons. The slides, videos, and *conversations* were viewed 467, 205, and 1479 numbers of times, respectively, over all modules, of which nearly 17 %, 20 %, and 20 % instances resulted in *likes* or *dislikes* feedback (Fig. 5A, B, C). We found that the feedback included more *likes* than *dislikes* for slides (8.0 ± 2.30 likes versus 0 ± 0 dislikes, Wilcoxon W = 45, n = 10 slides, p = 3.9 × 10^−3^; Fig. 5D), videos (7.4 ± 1.51 likes versus 0.60 ± 0.54 dislikes, W = 15, n = 5 videos, p = 0.063; Fig. 5E), and *conversations* (1.88 ± 0.24 likes versus 0.12 ± 0.033 dislikes, W = 6015, n = 149 *conversations*, p = 1.93 × 10^−19^; Fig. 5F). Participants also had the option to provide descriptive feedback on these elements. The subjective feedback was mostly positive, with participants frequently mentioning that they liked the given examples; one participant mentioned that they would have preferred to type their own answers in *conversations* (instead of choosing from pre-written text options). A word cloud created from the collated subjective feedback showed that the most frequently used words in feedback included “given”, “example”, “liked”, and “idea” (Fig. 5G). TreadWill allowed participants to revisit previously completed *conversations* to refresh their memory – this option was used 17 times by the participants.

**Figure 5:**
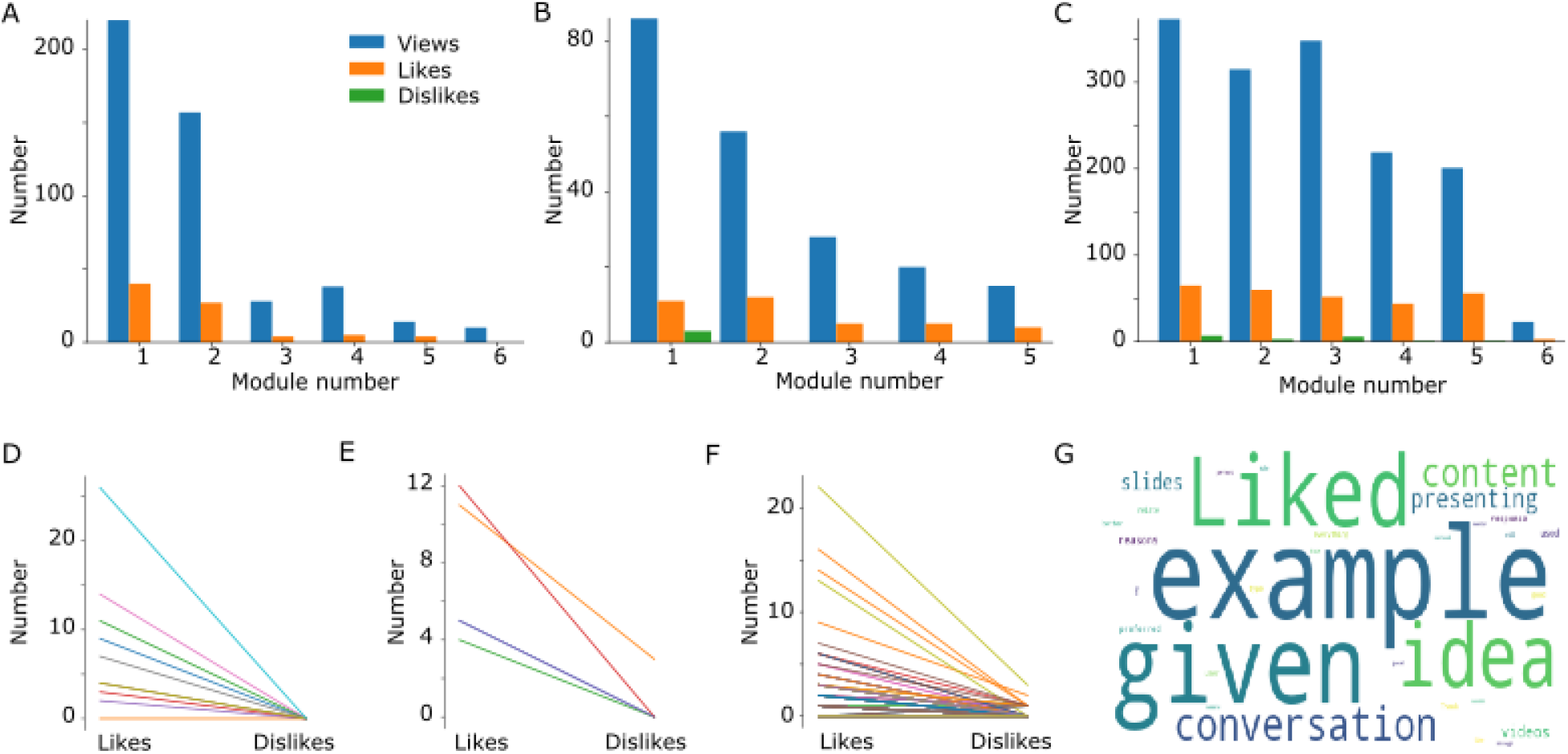
Feedback on the content. **A**,**B**,**C**, The bar graphs show the total number of views, likes, and dislikes received from the participants on the slides (**A**), the videos (**B**), and the conversations (**C**) presented in each module of TreadWill. There were no videos in Module 6. **D**,**E**,**F**, Slope graphs show that there were typically more likes than dislikes for slides (**D**, n = 10), videos (**E**, n = 5), and conversations (**F**, n = 149) presented in TreadWill. Each line indicates one slide, video, or conversation; some of the lines are overlapping. **G**, Word cloud of the descriptive feedbacks received from the participants on slides, videos, and conversations.

### Exploratory analysis: Patterns in participants’ activities

TreadWill allowed participants to attach one or more word tags, from a list of 44 tags, to posts in the *SupportGroup*. A word cloud of the tags used during the study revealed the topics most commonly discussed by the participants: “wasting time”, “loneliness”, “guilt”, “self-esteem”, and “trust” (Supplementary Fig. S5A). Next, we analyzed the entries made by the participants in the CBT forms (worksheets) to identify the common themes in their activities and concerns (see Supplementary Table S3 for brief descriptions of the forms). In the *Schedule activity worksheet*, the activities most frequently planned by the participants were related to completing a task, preparing or studying for an exam, exercising, sleeping on time, and waking up early (Supplementary Fig. S5B). The data from all other forms were tagged manually to homogenize the language used; a word cloud revealed that the major reasons for using the forms were “self-esteem”, “shame”, “relationship”, “anxiety, “failure”, “college”, and “competition” (Supplementary Fig. S5C). We checked the most commonly selected situations, thoughts, and beliefs from the lists presented to the experimental group participants. The most selected situation, thought, and belief were *“I am concerned about my career,”* “*I should be doing something better*,” and “*If I don’t work very hard, I’ll fail*,” respectively. (See Supplementary Fig. S6A, B, C for the top 10 selected situations, thoughts, and beliefs.)

## Discussion

We have presented the design of an unguided cCBT intervention – TreadWill – aimed at high user-engagement and universal accessibility. A fully remote randomized controlled trial with 598 participants was performed to test the effectiveness of TreadWill in reducing depression-related and anxiety-related symptoms. The results of the trial show that the full-featured (experimental) and the plain-text (active control) versions of TreadWill effectively reduced both PHQ-9 and GAD-7 scores compared with the waitlist control group, for the participants who completed at least three modules. Nearly 3 times more participants completed at least three modules in the experimental group than in the active control group. The extra features included in the experimental version increased adherence compared to the active control version, in terms of both the time of engagement and the number of modules completed. The results also show that the number of modules completed was correlated with the reduction in the symptom severity of a participant.

Two automated surveys presented during the intervention for taking participant feedback showed that the participants perceived TreadWill as useful and easy to use and found most of the interactive features helpful. Additionally, the feedback provided by the participants using *like* and *dislike* buttons on different elements of modules indicated that participants found the content relatable and useful.

We did not require a clinical diagnosis of depression for including participants in the study as our goal was to create an accessible tool catering to both clinical and sub-clinical populations. Given that the prevalence of sub-clinical depression, defined as a score in the range 5-9 in PHQ-9, is fairly high at 15-20% (Khaled, 2019; Kroenke et al., 2001, 2009), an unguided intervention can be immensely beneficial. We used only self-reported assessments for measuring symptom severity. While self-reported assessments have their drawbacks (Sato & Kawahara, 2011), it was essential given the pragmatic nature of the study with an unguided intervention. For the same reason, we also allowed participants undergoing other treatments (7% of our participants, see Table 1). We used only one questionnaire each for assessing depression and anxiety symptom severity. This decision was made keeping in mind that filling long questionnaires online is not a pleasant experience for the users and might increase dropout (Morriss et al., 2021). Also, while including multiple questionnaires for assessing the same disorder might improve validity, it also increases the risk of getting false-positive results by chance.

### Adherence rates in cCBT interventions

MoodGYM, one of the most popular cCBT intervention has reported moderate adherence rates of about 7% (Christensen et al., 2006) and 11.1% (Neil et al., 2009) and full adherence rates of 2.8% (Neil et al., 2009), 10% (Powell et al., 2013), and 15.8% (Farrer et al., 2011). Deprexis, another well-evaluated intervention, reported a full adherence rate of 7.5% in a fully unguided evaluation (Meyer et al., 2009).The high adherence rates observed in the trial settings often fail to transfer to the real world (Fleming et al., 2018). In real-world studies, adherence can be very low: 5.6% in Lara et al. (Lara et al., 2014), 13.11% in Morgan et al. (Morgan et al., 2017), and 5% in March et al. (March et al., 2021). In a fully remote trial of an app-based intervention, Arean et al. (Arean et al., 2016) reported that 57.9% of the participants did not even download their assigned apps. Similarly, in another study involving no human contact, Morriss et al. (Morriss et al., 2021) reported that only 57.3% of participants randomized to the experimental group signed up for the intervention and only 42.5% accessed it more than once. Morriss et al. (Morriss et al., 2021) further reported an attrition rate of 84.9% at 3-week follow-up with the attrition rate increasing at later follow-up points. In another recent study, Oehler et al. (Oehler et al., 2021) observed that the minimal dose was received by only 2.10% of the participants for the unguided version of iFightDepression. The adherence rate observed for TreadWill, 12.1% for moderate usage and 5.5% for full completion, is comparable to these real-world studies. At this adherence level, TreadWill can benefit a significant number of people from the general population as a fully automated and scalable intervention.

The 12.1% adherence rate for moderate usage was observed in our study despite additional challenges compared to other studies. Every step in our study, from participant recruitment to assessment, was fully automated; the lack of human contact is known to affect the commitment of the participants (Morriss et al., 2021). Further, it has been reported that male gender and young age significantly increase the chance of dropout (Karyotaki et al., 2015). The average age of our participant group was 23.5 years, and 79% were male, which could have contributed to the dropout. Our participants were non-native English speakers in an LMIC (the intervention was developed in English to ensure future scalability in other countries). Contrary to the practice of giving money or gift cards to participants (Arean et al., 2016; Clarke et al., 2009; Pratap et al., 2018; Schure et al., 2019), we did not reward participants for submitting assessments or for participation. In several studies (Arean et al., 2016; Clarke et al., 2009; Pratap et al., 2018; Schure et al., 2019), the participants were paid even after submitting the baseline assessments. The practice of paying participants is likely to influence adherence to the intervention due to the rule of reciprocity (Cialdini, 1987) and to influence the assessment responses. Participants getting paid might feel that they owe it to the researchers to use the program and try to give answers in the assessments they think the researchers expect. Not giving a reward also supports our pragmatic trial design, as in the real world paying participants to use the intervention will be unsustainable.

### Implications

Our study shows that even in low-resource settings, cCBT based interventions can help without any expert support. This implication is immensely encouraging as the number of mental health professionals is extremely low in India (Patel et al., 2016; but see Garg et al., 2019). Our study establishes TreadWill as a potential population-level intervention. This is among the largest studies conducted in India on digital mental health (Kanuri et al., 2020; Mehrotra et al., 2018; Rodriguez-Villa et al., 2021; Srivastava et al., 2020). Our study is also the first fully online trial conducted in India and provides a template for conducting online trials for other mental health conditions in the country. Future work should focus on strategies like — using gamification, using serious games, or using a chatbot to build therapeutic alliance — to improve adherence to self-help interventions.

## Supporting information

CONSORT Checklist

## Data Availability

The de-identified data analyzed in the current study are available from the corresponding author on reasonable request.

## Acknowledgements

We thank Romit Chaudhary, Divya Chauhan, Vinay Agarwal, Sahars Kumar, Pearl Sikka, Rahul Gupta, Nikhil Vanjani, and Sandarsh Pandey for their help in development of TreadWill. We thank Pranjul Singh, Aditya Patil, and Pearl Sikka for their help in developing the videos of TreadWill. We thank Braj Bhushan, Shoukkathali K, Rita Singh, Akanksha Awasthy, and Gitanjali Narayanan for helpful discussions. We thank Dr. Shikha Jain, Mrityunjay Bhargava, Pratibha Mishra, Jagriti Agnihotri, Swastika Tandon, Akash A., Harsh Agarwal and IIT Kanpur media cell for promoting the visibility of the trial. We thank Silky Gupta and Aarush Mohit Mittal for help with data analysis and feedback on the manuscript. We thank members of the Lab of Neural Systems for testing the beta version of TreadWill and giving feedback on the content and user experience. This work was supported by the Cognitive Science Research Initiative of the Department of Science & Technology [grant number DST/CSRI/2018/102]. The funding agency had no role in the design or implementation of the study and in the interpretation of the results.

## Author contributions

AG and NG conceptualized the project; AG, RJC, SW, AB, and NG designed the research; AG, RJC, SW, PS, and KRK developed the intervention; AG, AB, and NG recruited participants; AG and NG analyzed data; AG and NG wrote the paper with inputs from all co-authors.

