## Supplement for "TreadWill: Development and pragmatic randomized controlled trial of an unguided, computerized cognitive behavioral therapy intervention in a lower middle-income country"

**Supplementary tables S1-S5.**

**Supplementary figures S1-S6.**

**Supplementary Table S1: Automated notifications**

| Purpose | Type | Frequency | Group |
|---|---|---|---|
| Notify the user to use the site if they have been inactive | Email | 3, 7, 14, 28, 56, and 89 days after the user's last activity time | Experimental group |
| Notify the user's family member to nudge the user to use the program | Email | 5, 10, 30, and 70 days after the user's last activity time | Experimental group |
| Weekly update on the user's progress to the user's family member | Email | Once a week if the user has been active in the last 15 days | Experimental group |
| Notification about scheduled activities | SMS | Once per task | Experimental group |
| Notify that the next module has been unlocked | Email | Once | Experimental group |
| Notify the user 10 minutes before their preferred login time | Email and SMS | Daily, if the user has been active in the last 7 days | Experimental group |
| Notify the user for the number of days left | Email | 30, 15, 7, and 3 days before the completion of the 90 days | Experimental group |
| Notify the user to fill the follow-up PHQ-9 and GAD-7 questionnaires | Email and SMS | 90, 94, 98, 105, and 120 days after:<br>• the last activity time if the user hasn't completed all the modules<br>• the time the user completed the final module | Experimental and Active control group |
| Congratulate the users who have completed all the 6 modules | Email | Once after all the modules are completed | Experimental and Active control group |
| Nudge the user to complete the registration process | Email | 1, 4, 8, 16, 32, and 64 days after they started the registration process | Experimental and Active control group |
| Notify the user that their 90 days period is over | Email | 90 days after the user visits the dashboard for the first time | Experimental and Active control group |
| Notify the user to complete the survey for users who have not completed the surveys | Email | 90, 94, 98, 105, and 120 days after the user visits the dashboard for the first time | Experimental and Active control group |

| | | | |
|---|---|---|---|
| Nudge the user to complete the first baseline GAD-7 questionnaire | Email | 1, 3, and 7 days after they submitted the baseline PHQ-9 | Waitlist control group |
| Notify the waitlist group user that the experimental group intervention is now unlocked | Email | Once after 72 days after they have been randomized | Waitlist control group |
| Notify the waitlist group user to fill the PHQ-9 and GAD-7 questionnaires after the 42-day interval | Email and SMS | 42, 43, 45, 48, 52, 57, and 72 days after randomization | Waitlist control group |

**Supplementary Table S2: Modules in TreadWill**

| Number | Module Name | Description |
|---|---|---|
| 1 | Basics | The first module provided psychoeducation about CBT, depression, and generalized anxiety disorder. This module oriented the participant towards CBT and addressed common concerns about the disorders among patients. |
| 2 | Behavioral Activation | The second module provided the rationale for behavioral activation and helped the participants develop a balanced schedule. The goal was to provide mastery and pleasurable activities, reduce rumination, and increase socialization. |
| 3 | Identify NATs | This module aimed to help the participants develop skills to identify negative automatic thoughts (NATs). The participants learned different techniques like "Visualizing the situation," "Heightening physiological response," and others. They also learned how to apply one or multiple techniques to identify automatic thoughts. |
| 4 | Challenge NATs | The goal of this module was to teach techniques to evaluate and challenge their negative automatic thoughts. The participants learned skills like "Identifying cognitive distortions" and "Socratic questioning" to evaluate and challenge their negative automatic thoughts. They also learned about *problem solving and acceptance* in case their negative automatic thoughts were true. |
| 5 | Modifying Beliefs | This module taught the participant how to modify the more deeply set intermediate and core beliefs. This module ensured that the intervention had a prolonged effect on the participant. |
| 6 | Staying Happy | This module warned the participant that they were going to face setbacks in the future. The module advised the participant to regularly exercise the skills developed in the program to prevent another episode of depression. |

**Supplementary Table S3: CBT worksheets included in TreadWill**

| Worksheet name | Description |
|---|---|
| Thought record worksheet | This form allowed the users to write a situation and the corresponding negative automatic thought and emotion. It provided all the techniques to evaluate and challenge this negative automatic thought. |
| Core belief worksheet | This form allowed the users to evaluate their old maladaptive beliefs that they wanted to modify and new adaptive beliefs that they wanted to adopt. |
| Behavioral experiment worksheet | This form allowed the users to design a behavioral experiment to evaluate a maladaptive belief. |
| Problem solving worksheet | This form allowed the users to brainstorm potential solutions to a problem, identify the best one, and implement it. |
| Prepare for setback worksheet | This form allowed the users to think of potential setbacks and adaptive ways to respond to the setbacks. |
| Schedule activity worksheet | This form had two independent parts. In part 1, the users had the option to write a mastery activity that they would like to complete. In part 2, the users could write an activity that they would like to do and the time they would like to do it. This activity could be a recurring activity (e.g. – exercising) or a one-time activity (e.g. – make an important call). The experimental group users could opt to receive an SMS notification 10 minutes before the start of the activity. |

**Supplementary Table S4: Survey questions**

Each question included a statement, to which the participant responded using a 5-point Likert scale: 2 = strongly agree, 1 = somewhat agree, 0 = neither agree nor disagree, -1 = somewhat disagree, -2 = strongly disagree.

| Question Number | Question in the experimental group | Question in the active control group |
|---|---|---|
| 1 | TreadWill has positively affected my life. | TreadWill has positively affected my life. |
| 2 | TreadWill was easy to navigate. | TreadWill was easy to navigate. |
| 3 | I found the language used in TreadWill easy to understand. | I found the language used in TreadWill easy to understand. |
| 4 | I found the examples relatable. | I found the examples relatable. |
| 5 | I found the Forms easy to use. | I found the Forms easy to use. |
| 6 | I found conversations engaging. | I found the discussions engaging. |
| 7 | The SupportGroup was a useful component of the program. | I would prefer to have a way to post my queries in the program. |
| 8 | The PeerGroup was a useful component of the program. | I would prefer to have a way to interact with other users in the program. |
| 9 | Involving the friend with me in the program helped me stay motivated. | I would prefer to involve a friend with me in the program to help me stay motivated. |
| 10 | I found the email reminders helpful. | I would prefer to have email reminders. |
| 11 | I found the SMS reminders helpful. | I would prefer to have SMS reminders. |
| 12 | I found the Learning videos helpful. | I would prefer Learning through videos than plain text. |
| 13 | I found Learning slides helpful. | I would prefer Learning through slides than plain text. |
| 14 | I found the game elements (Score, Find the friendly face etc.) helpful. | I would have preferred to have some game elements in the program. |
| 15 | Acting as a therapist to an apprentice helped me take a more objective view of my problems. | I think that acting as a therapist to an apprentice would have helped me take a more objective view of my problems. |

**Supplementary Table S5: Characteristics of completers and non-completers**

Baseline and demographic characteristics of the participants who completed all modules (completers) versus the participants who did not complete all modules (non-completers). There was a single completer in the active control group and has been pooled with the 10 participants of the experimental group in the table. There is no significant difference in age or baseline PHQ-9 scores between completers and non-completers.

| **Group (n)** | **Completers (n=11)** | **Non-completers (n=382)** |
|---|---|---|
| **Age**, Mean (SE) | 25.09 (1.301) | 23.56 (0.21) |
| **Gender**, (male, female) | (8, 3) | (303, 79) |
| **Traumatic event or death of a loved one**, (Yes, No) | (0, 11) | (40, 342) |
| **Joining for help**, Yes/No | (11, 0) | (326, 56) |
| **Secondary help**, (None, Counselling, Medication, Both) | (10, 0, 1, 0) | (353, 9, 15, 5) |
| **Occupation**, (High School Student, Between school and college, College student, Coaching after college, Working professionals, Self employed, Freelancers, Volunteers) | (0, 0, 4, 4, 2, 1, 0, 0) | (3, 12, 212, 62, 77, 10, 5, 1) |
| **PHQ-9**, Mean (SE) | 10.82 (1.10) | 10.79 (0.18) |

**Supplementary Figure S1**

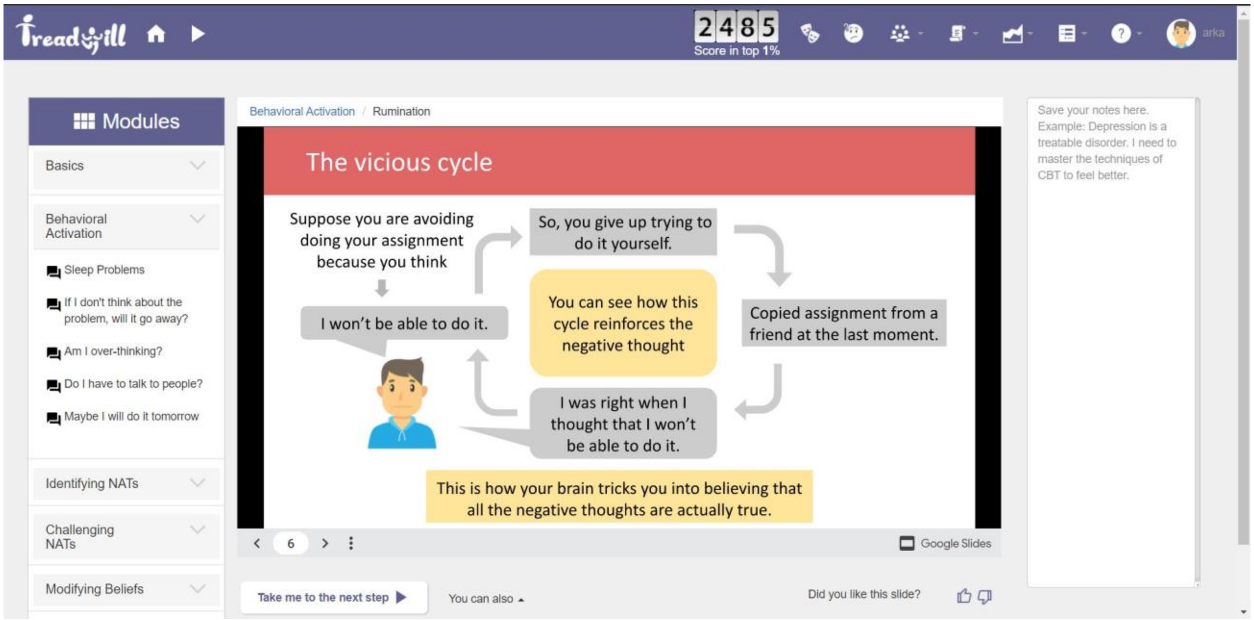


**Supplementary Figure S1**. Snapshot of a slide in TreadWill. To the left of the slide is a side-bar for quickly accessing the other modules. To the right of the slide is a text-box for the user to take notes. Below the slide is a button to access the next step and to give feedback on the slide.

**Supplementary Figure S2**

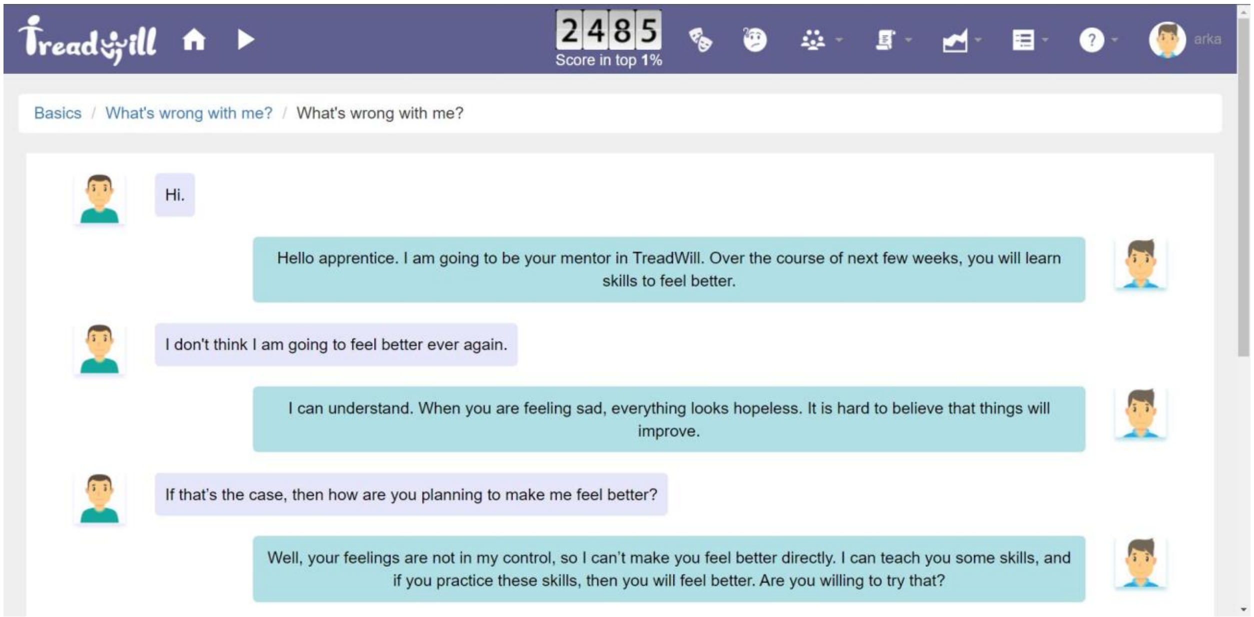


**Supplementary Figure S2**. Snapshot of an ongoing *conversation* between the participant (right) and an automated virtual patient (left), that was tailored to have a similar profile as the participant. The participant practiced the CBT techniques while trying to help the simulated patient.

**Supplementary Figure S3**

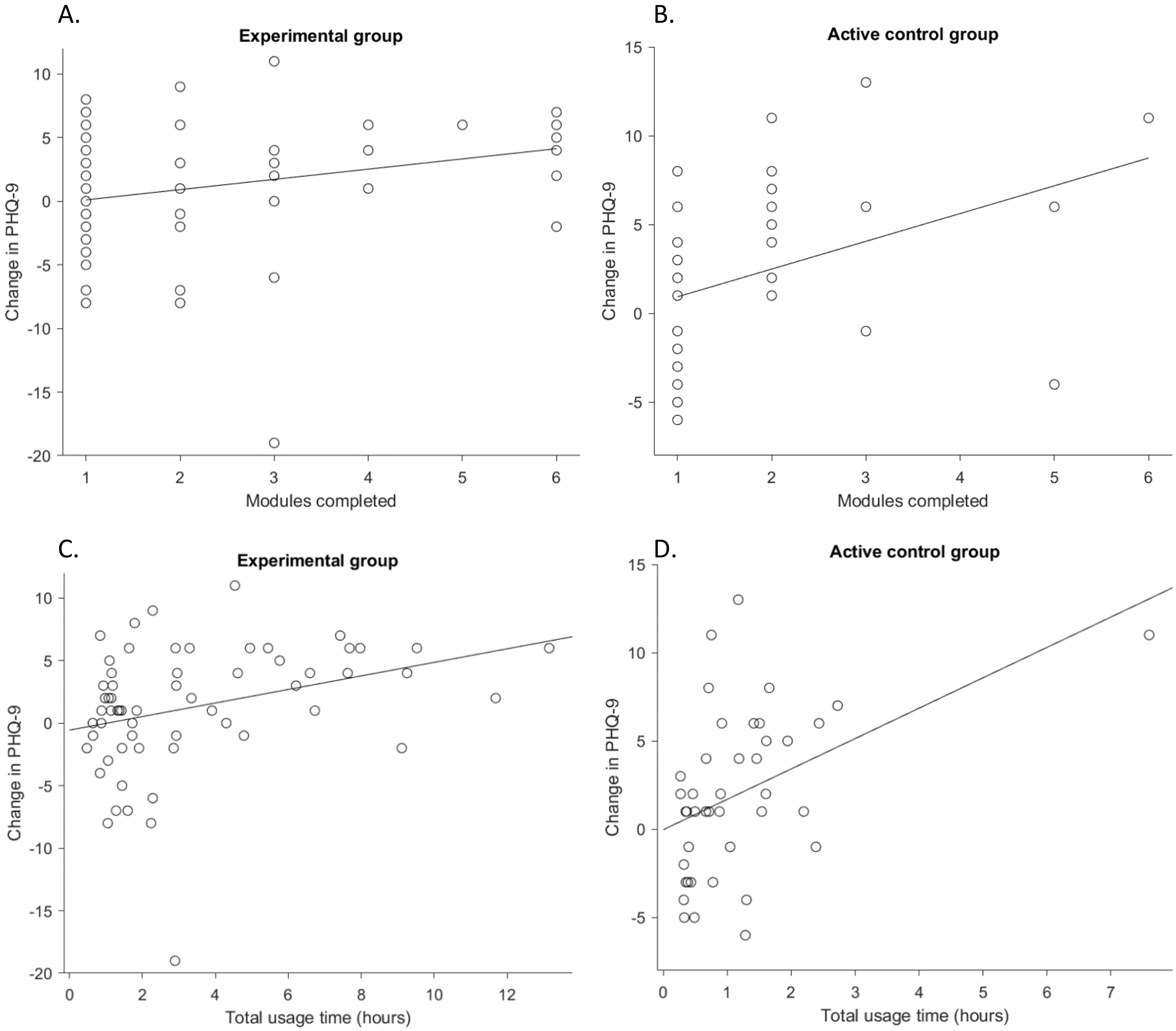


**Supplementary Figure S3**. Scatter plots show correlations between the usage of TreadWill and the reduction in depressive symptoms. **A,B**, Reduction in PHQ-9 score versus the number of modules completed by the experimental group participants (A) and the active control group participants (B). **C,D,** Reduction in PHQ-9 score versus the total usage time in hours for the experimental group participants (C) and the active control group participants (D). In all cases, the reduction in PHQ-9 score was calculated by subtracting the last PHQ-9 score (excluding follow-up) from the baseline score. Some points in the graphs are overlapping.

**Supplementary Figure S4**

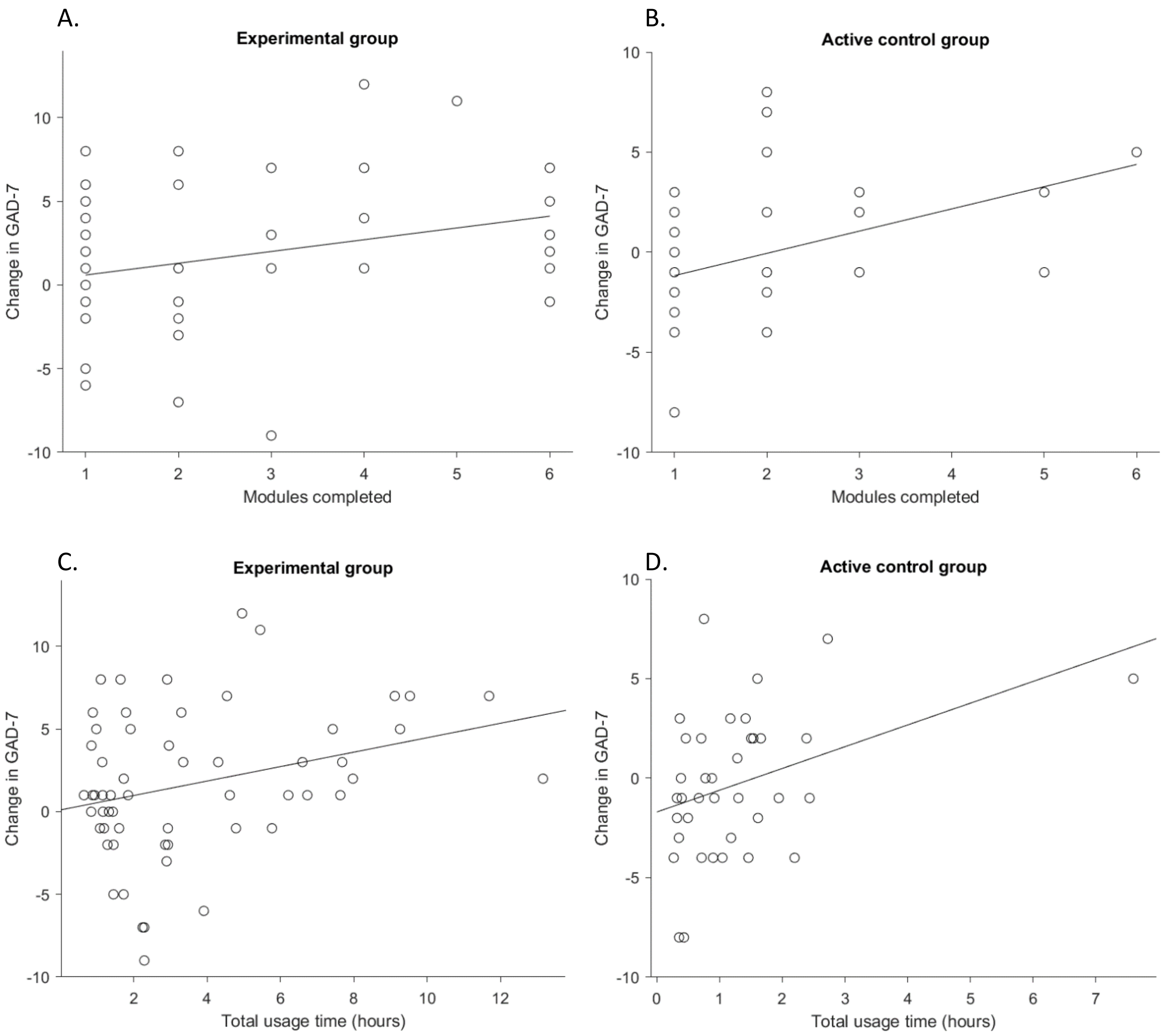


**Supplementary Figure S4**. Scatter plots show correlations between the usage of TreadWill and the reduction in anxiety symptoms. **A,B**, Reduction in GAD-7 score versus the number of modules completed by the experimental group participants (A) and the active control group participants (B). **C,D,** Reduction in GAD-7 score versus the total usage time in hours for the experimental group participants (C) and the active control group participants (D). In all cases, the reduction in GAD-7 score was calculated by subtracting the last GAD-7 score (excluding follow-up) from the baseline score. Some points in the graphs are overlapping.

**Supplementary Figure S5**

A.

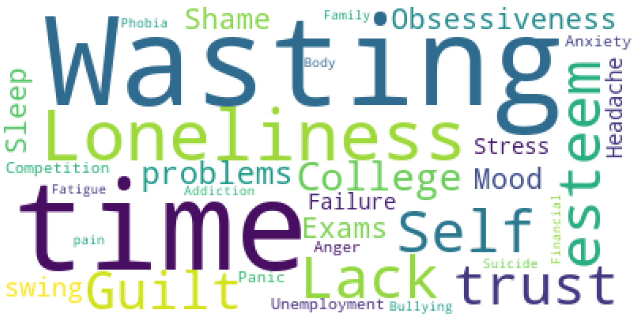


B.

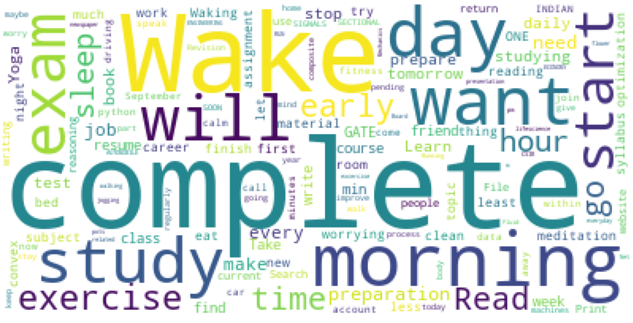


C.

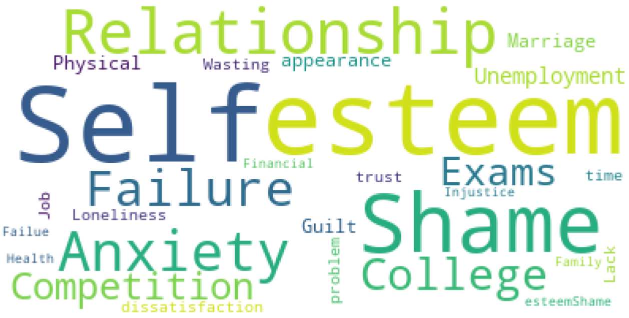


**Supplementary Figure S5**. Words frequently used by the participants. **A**, A word cloud of tags used in the *SupportGroup* posts by the participants. **B**, A word cloud of activities set by the participants in the *Schedule activity worksheet*. **C**, A word cloud of tags representing the problems mentioned by the participants while using the CBT forms (worksheets). This word cloud combines the data from the *Thought record worksheet, Core belief worksheet, Behavioral experiment worksheet, Problem solving worksheet,* and *Prepare for setback worksheet*. The worksheets were tagged by us to standardize the language used. Panel A includes data from the experimental group (active control group participants did not have access to the *SupportGroup*), while panels B and C include data from both the experimental and the active control group participants.

**Supplementary Figure S6**

**A.**

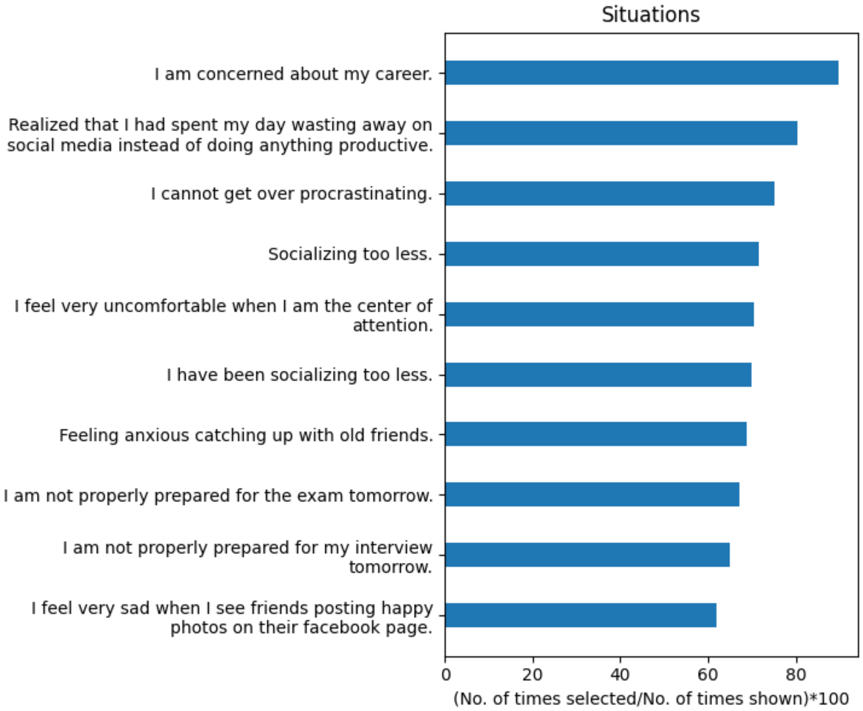


**B.**

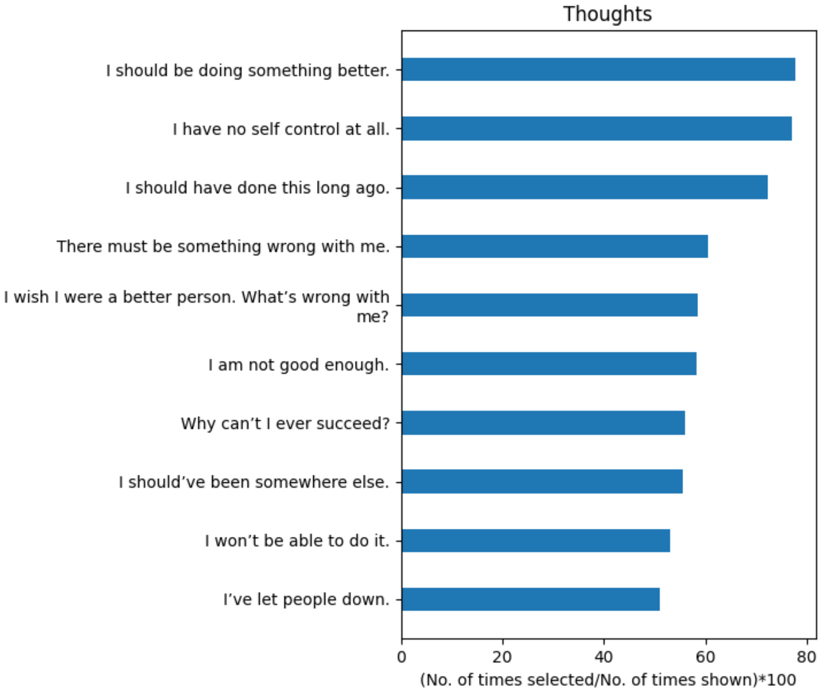

**C.**

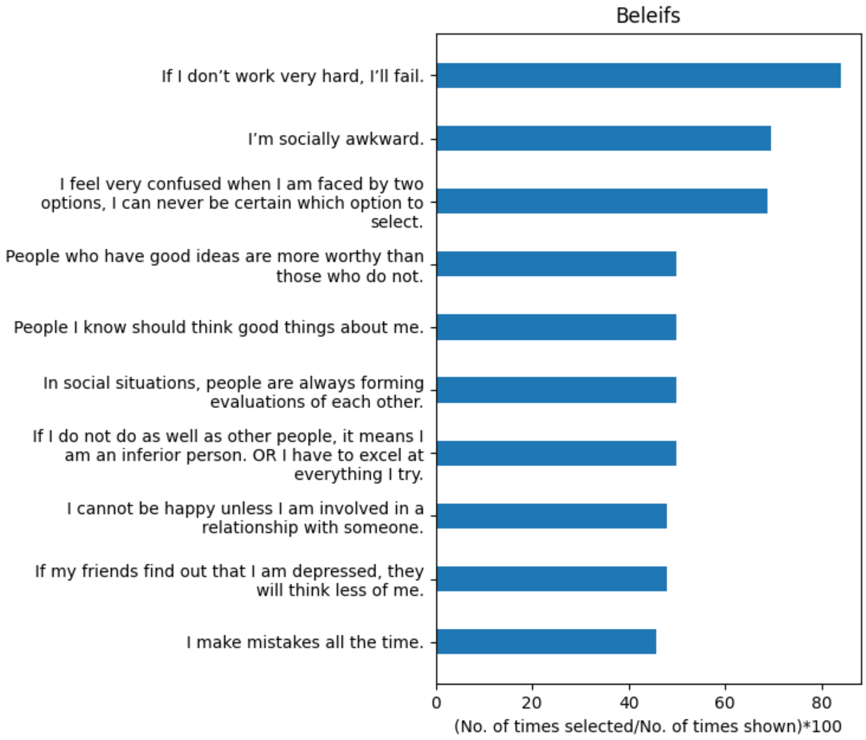


**Supplementary Figure S6**. The bar graph shows the 10 most frequently selected situations (A), thoughts (B), and beliefs (C) by the experimental group participants. The participants selected these from a list containing 2 to 6 beliefs, thoughts, and situations presented to them when they clicked on the *Discuss* cards for the first time. The total number of beliefs, thoughts, and situations shown was 71 for high school students, 84 for college students, and 60 for working professionals.
